# Characterizing the transmission patterns of seasonal influenza in Italy: lessons from the last decade

**DOI:** 10.1101/2020.11.29.20240457

**Authors:** Filippo Trentini, Elena Pariani, Antonino Bella, Giulio Diurno, Lucia Crottogini, Caterina Rizzo, Stefano Merler, Marco Ajelli

## Abstract

**Background:** Despite thousands of influenza cases annually recorded by surveillance systems around the globe, estimating the transmission patterns of seasonal influenza is challenging.

**Methods:** We develop an age-structured mathematical model to influenza transmission to analyze ten consecutive seasons (from 2010-2011 to 2019-2020) of influenza epidemiological and virological data reported to the Italian surveillance system.

**Results:** We estimate that 18.4%-29.3% of influenza infections are detected by the surveillance system. Influenza infection attack rate varied between 18.0% and 35.6% and is generally larger for seasons characterized by the circulation of A/H3N2 and/or B types/subtypes. Individuals aged 14 years or less are the most affected age-segment of the population, with A viruses especially affecting children aged 0-4 years. For all influenza types/subtypes, the mean effective reproduction number is estimated to be generally in the range 1.1-1.4 (8 out of 10 seasons) and never exceeding 1.55. The age-specific susceptibility to infection appears to be a type/subtype-specific feature.

**Conclusions:** The results presented in this study provide insights on type/subtype-specific transmission patterns of seasonal influenza that could be instrumental to fine-tune immunization strategies and non-pharmaceutical interventions aimed at limiting seasonal influenza spread and burden.

## INTRODUCTION

Annual influenza epidemics cause a marked excess of mortality and hospitalization as well as significant economic and healthcare burden [1]. Worldwide, influenza-associated respiratory deaths are estimated to be in the range 4.0-8.8 per 100,000 individuals, with an impressive peak of 17.9-223.5 deaths per 100,000 people for individuals aged 75 years or more [2]. To monitor influenza spread, surveillance is carried out worldwide by means of records of patients showing influenza-like illness (ILI) and virological investigation of circulating types/subtypes. Moreover, due to the continuous antigenic changes of the virus, the investigation of the circulating influenza types and subtypes and the assessment of their burden in specific age classes across different seasons is crucial to update the composition of the vaccine and increase its efficacy [3]. However, due to the low proportion of individuals developing clinical symptoms after influenza infection [4] and thus the low proportion of individuals consulting general practitioners [5], the picture returned by the surveillance data is far from being representative of the true influenza burden.

Mathematical models of infectious disease transmission represent key tools to properly interpret the observed data and to provide quantitative estimates of quantities that hard to measure directly [6-9]. In this study, we use mathematical modeling to estimate three key epidemiological indicators: i) the influenza infection attack rate (overall and by age), which corresponds to the proportion of individuals infected by influenza over the entire course of the season; ii) the effective reproductive number, Re - i.e., the number of secondary infections caused by an influenza infectious individual; and iii) the age-specific susceptibility to infection by virus type/subtype. Each epidemiological indicator is estimated for ten influenza seasons (from 2010-2011 to 2019-2020) in Italy and for each circulating influenza type/subtype.

Modeling and comparing the influenza types/subtypes allows us elucidating type/subtype-specific features and to what extent the co-circulation of different types/subtypes alters the transmission patterns at the population level of the single type/subtype. The estimates provided in this study shed new light on the transmission dynamics of seasonal influenza, showing that the total infection attack rate has a low variability across different seasons, the reproduction number is not markedly different by influenza type/subtype, and underage individuals play a central role in spreading the infection.

## METHODS

### Influenza like illness and virological surveillance data

We analyze the data reported to the Italian epidemiological and virological influenza surveillance system from 2010-2011 to 2019-2020. Briefly, general practitioners (GPs) and pediatricians (PDs) are asked to report weekly influenza like illness cases (ILI, defined as acute onset of fever > 38°C, + respiratory symptoms+ one of these symptoms: headache, general discomfort, asthenia) occurring during the year, from week 42 to week 17, using standardized forms. Specific information regarding age (0-4, 5-14, 15-64, >64 years) and influenza vaccine status are collected and reported using web-based electronic Case Report Forms [10]. It is important to highlight that GPs and PDs are also able to define their assisted population by age group because every individual in Italy has to be appointed to a specific GP through the regional health service. This measure allows calculating the incidence of ILI cases by age group at the National and regional level (every year 2% of the regional population is requested to be under surveillance as per indication of the National Public Health Institute that coordinates the surveillance scheme).

### Virological surveillance data

For surveillance of circulating influenza viruses – sampling kits are sent out to regional coordinator for surveillance that randomly select 1 GP per week between week 46 and week 10 to collect throat swabs of the first ILI-patients seen. These swabs are analyzed at the regional Reference Laboratories distributed in 15 different Italian regions [11]. Results are collected and reported using web-based electronic CFR from the National Influenza Centre (NIC). Every season are collected approximately 2,000 samples with a proportion of positive specimens of about 34% [12].

Virological data from Italian region of Lombardy for the seasons from 2010-2011 to 2016-2017 season [13] and from the National Influenza Surveillance Scheme from 2017-2018 to 2019-2020 season [14] are used in our analysis. A comparison between national virological surveillance and virological data for Italian region of Lombardy from 2010-2011 to 2016-2017 season is reported in the Appendix [see Additional File 1: pp 4].

### Seroepidemiological data

We analyze age-specific data about the 2009 A/H1N1 influenza pandemic. Briefly, we analyze seroepidemiological data collected before and at the end of the 2009 A/H1N1 pandemic to assess both the pre- [15] and post- [16] pandemic susceptibility to infection and the level of immunity by age-group to the 2009 A/H1N1 influenza virus in the Italian general population. The level of immunity against the 2009 A/H1N1 influenza virus in pre- and post-pandemic sera are determined using left over sera taken for diagnostic purposes or routine ascertainment obtained from clinical laboratories. The antibody titres are measured by the haemagglutination inhibition (HI) assay (the presence of protective antibody (≥1⍰40), are calculated using exact binomial 95% CI on both pre- and post-pandemic serological data) [16].

For each season we consider two datasets: i) the weekly incidence of ILI cases by age group (four age groups: 0-4 years, 5-14 years, 15-64 years, and 65+ years) [17], and ii) the share of samples collected among ILI cases testing positive for each of three influenza types/subtypes (namely, A/H1N1pdm09, A/H3N2, and B – both Victoria and Yamagata) by age (same four age groups of the ILI cases) [13,14] and the seroepidemiological profile of the Italian population against 2009 H1N1 influenza pandemic.

In addition to the epidemiological data, for each season, we collect data about the influenza vaccination coverage by age group [18] and age structure of the Italian population [19]. Estimates of vaccine effectiveness by type/subtype are taken from Belongia et al. [20]. Details on the data used for the analyses are presented in the Appendix [see Additional File 1: pp 2-3].

### Estimation of ILI reporting rate

We subtract the proportion of samples positive for 2009 A/H1N1 pandemic virus (A/California/07/2009) identified after and before the 2009 A/H1N1 pandemic virus circulated in Italy to obtain an estimate of the infection attack rate (AR) of the pandemic for individuals aged 0-14 years and 15+ years. We label these quantities as *AR*_0-14_ and *AR*_15+_, respectively. We then sum the weekly ILI cases reported to the surveillance system during the pandemic in the same two age groups and multiply it by the share of samples collected among ILI cases testing positive for A/H1N1pdm09. These quantities, denoted as 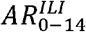 and 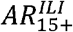 essentially represent the attack rate detected by the Italian surveillance system. Therefore, we can estimate the reporting rate of the surveillance system for the age group 0-14 years and 15+ years as the ratios 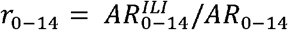 and 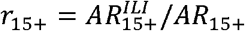, respectively. Details are provided in the Appendix [see Additional File 1: pp 4-5].

### Estimation of age-specific infection attack rates for seasonal influenza

We leverage the estimated reporting rates by age of the surveillance system to estimate the infection attack rate of each type/subtype for each of the analyzed influenza seasons. In particular, we define *ILI*_*a*_(*w; y*) the incidence of *ILI* cases reported to the surveillance system for age group a on week *w* of season *y*. We also define *f*_*a*_ (*y, s*) the share of *ILI* samples for age group a testing positive for type/subtype *s* in season *y*. Therefore, following the procedure presented in [16], we can estimate the incidence of influenza cases linked to type/subtype *s* for each age group and influenza season as 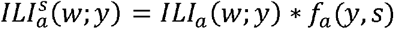. By summing over all weeks, we can get an estimate to the age-specific AR detected by the surveillance system. Using the reporting rate by age of the surveillance system (see previous section), we can estimate the influenza infection AR by age in season *y* as 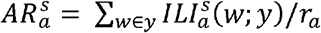 Note that, as we have estimates of the reporting rate for only two age groups (0-14 and 15+ years) but ILI cases for four age groups (0-4, 5-14, 15-64, 65+ years), we apply *r*_0-14_ to age groups 0-4 and 5-14 years and we apply *r*_15+_ to age groups 15-64 and 65+ years. Details are provided in the Appendix [see Additional File 1: pp 4-5].

### Modeling analysis

To obtain posterior estimates of the epidemic reproduction number and age-specific susceptibility to infection for each influenza type/subtype and season, we use a Bayesian approach. First, we define an ordinary differential equation influenza transmission model following the classic SLIR scheme. Essentially, susceptible individuals (S) can acquire the infection and enter the latent compartment (L) after contact with an infectious individual (I). After a latent period, latent individuals become infectious and can transmit the infection. Finally, after an infectious period, infectious individual recovers and enter the removed (R) compartment. The latent period is set to 1.5 days [21] and the infectious period is set to 1.2 days in such a way that the resulting generation time is 2.7 days, in agreement with the literature [22].

The population is further divided into 86 age groups (1-year age groups from 0 to 84 year and one age group for individuals aged 85 years or older) to account for a heterogeneous contact pattern by age, which is well known to be a major determinant of influenza dynamics [23,24]. In particular, we use the age-specific contact matrix derived for the Italian population at 1-year age resolution presented in [24]. In addition, the model accounts for the susceptibility to infection by age group (four age groups: same as those of the ILI data), that captures social, hygienic, and biological determinants (e.g., residual immunity to the circulating type/subtype) of the infection, which are not captured by the heterogeneous age-mixing pattern [7,16,25,26,27]. We set the initial condition of the system by considering the observed age-specific vaccination coverage and type/subtype-specific vaccine efficacy [20]. The model is regulated by four free parameters: the transmission rate, and the susceptibility to infection of age groups 5-14, 15-64, 65+ years relative to the age group 0-4 year (for which the susceptibility to infection is set to the reference value of 1).

For each season and influenza type/subtype, we explore the likelihood of observing the estimated type/subtype-specific infection attack rate 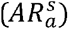 given a set of model parameters by using a differential evolution Markov chain Monte Carlo (MCMC) approach [28]. To address potential bias due to paucity of data that may also affect the goodness of MCMC convergence, we report results only for the types/subtypes accounting for at least 15% of the samples testing positive for influenza. The posterior distribution of the (season- and type/subtype-specific) effective reproduction number R_e_ is then computed by using the next-generation matrix approach [29]. In particular, Re can be computed as the dominant eigenvalue of the Next Generation Matrix (NGM) [29] associated with the dynamical system considered, accounting not only for age patterns in the Italian contact matrix but also for the susceptibility to infection among different age classes. Details on the methodology are reported in the Appendix [see Additional File 1: pp 5-8].

## RESULTS

In each of the ten analyzed seasons, the incidence of ILI cases reported to the Italian surveillance system shows an annual epidemic characterized by a peak occurring in February with the exception of the 2016-2017 season when the peak was recorded in December (Fig. 1A). The maximum peak week incidence varied from 6.1 cases per 1,000 individuals in the 2015-2016 season to 14.7 cases per 1,000 individuals in the 2017-2018 season (Fig. 1A). The share of ILI cases testing positive for influenza ranges from 32.9% in the 2019-2020 season and 52.2% in the 2014-2015 season (Fig. 1B) [see also Additional File 1: pp 2-3]. All the analyzed seasons are characterized by the co-circulation of A/H1N1pdm09, A/H3N2, and B types/subtypes, although in some seasons most samples tested positive for one virus only (e.g., in the 2011-2012 season, 87% of the samples testing positive for influenza are associated with A/H3N2 infection), while in other seasons all the types/subtypes showed a similar share (e.g., in the 2014-2015 season) – see Fig. 1B.

**Figure 1.**
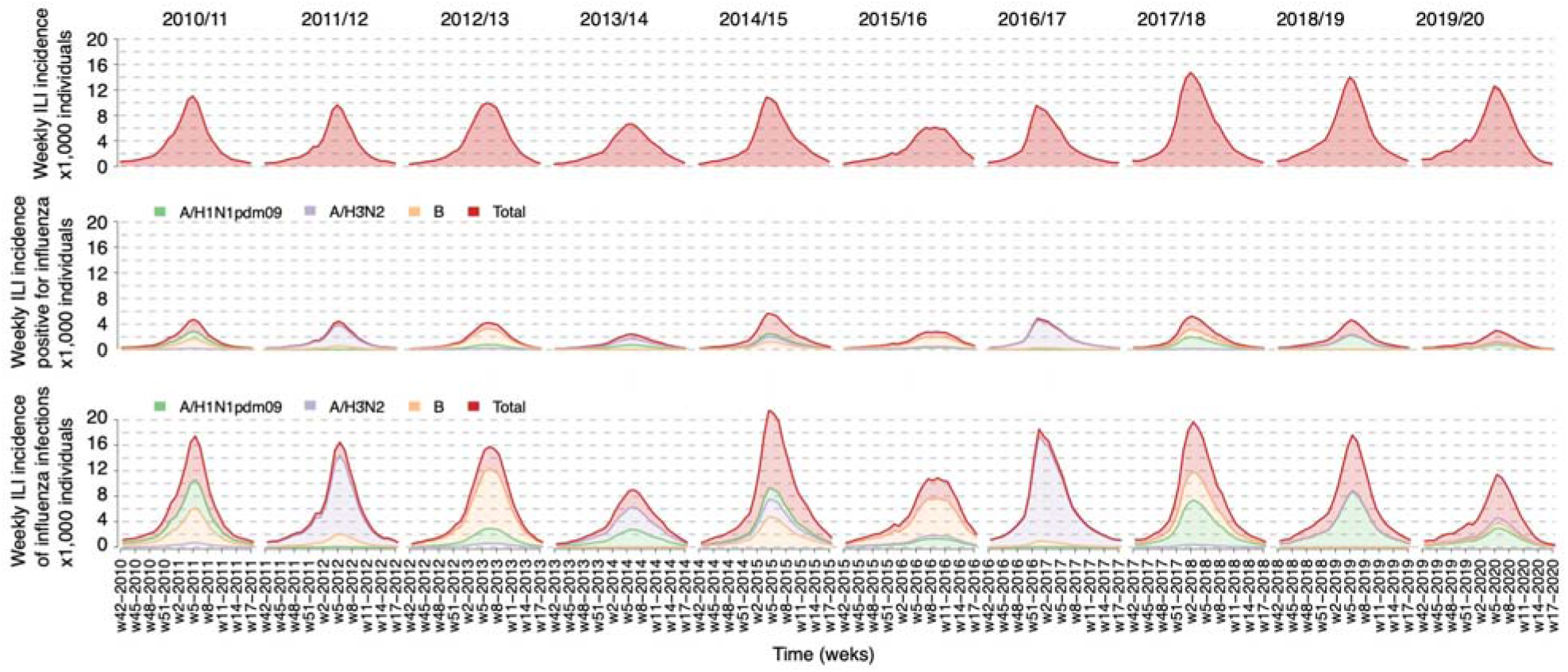
**A** Weekly incidence of ILI cases per 1,000 individuals reported to the Italian surveillance system for the ten seasons from 2010-2011 to 2019-2020. **B** As A, but for ILI cases testing positive for influenza virus, total and by type/subtype. **C** As B, but for the estimated incidence of influenza infections, total and by type/subtype, adjusted by considering the reporting rate.

The estimated reporting rates of the Italian surveillance system are *r*_0-14_ 0.184 (95%CI: 0.164-0.208) and *r*_15+_ 0.293 (95%CI: 0.204-0.445) for individuals aged 0-14 years and 15+ years, respectively. These values are in general agreement with independent estimates available in the literature [5, 25, 32]. When accounting for the reporting rate, we have a clearer picture of the actual magnitude of the seasonal influenza epidemics by type/subtype (Fig. 1C). The peak week incidences ranged from 9.0 influenza infections per 1,000 individuals (95%CI: 6.8-11.7 infections per 1,000 individuals) to 21.6 influenza infections per 1,000 individuals (95%CI: 16.3-28.1 infections per 1,000 individuals). For each season, the infection attack rate (all types/subtypes) ranged from 18.0% (95%CI: 14.5%-21.6%) in the 2013-2014 season to 35.6% (95%CI: 34.8%-36.4%) in the 2017-2018 season (Fig. 2). The type/subtype-specific infection attack rate is instead highly variable across seasons (Fig. 2). The largest infection attack rate for A/H3N2 influenza was 20.6% (95%CI: 17.1%-24.0%), observed during the 2016-2017 season; for B it was 24.7% (95%CI: 20.7%-28.3%) in the 2012-2013 season; for A/H1N1pdm09 it was 14% (95%CI: 13.5%-14.6%) in the 2017-2018 season. Interestingly, the second largest infection attack rate for A/H1N1pdm09 was recorded the season following the pandemic, 12.4% (95%CI: 9.8%-14.9%), and it was slightly lower than that observed during the pandemic year, 16.3% (95%CI: 9.4%-23.1%) [16]. Essentially no circulation of A/H1N1pdm09 was recorded in 2011-2012 and 2016-2017, while very low circulation was observed in the 2012-2013, 2013-2014 and 2015-2016 seasons (estimated infection attack rates lower than 5%).

**Figure 2.**
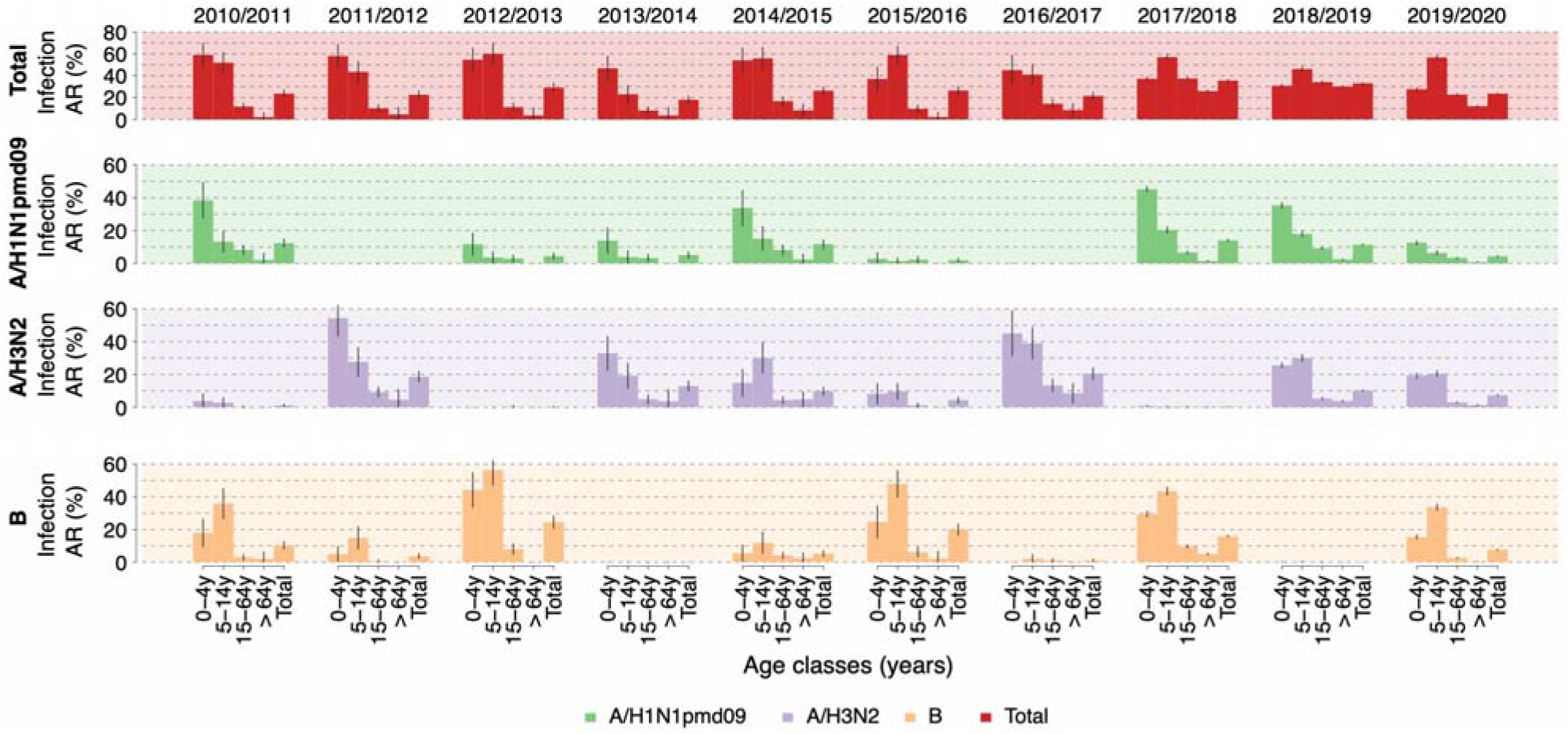
Estimated mean attack rate (total and by age group) for all types/subtypes (total) and by type/subtype in the ten seasons from 2010-2011 to 2019-2020. The vertical lines represent 95%CI calculated with an exact binomial test applied to the number of samples testing positive for each type/subtype and the number of tested samples.

By considering infection attack rates by age, the figure is very heterogeneous across different seasons and influenza subtype (Fig. 2). In particular, for A/H1N1 subtype, the infection attack rate decreases by age group, while for A/H3N2 this hold only when it is the predominant subtype of the season. During seasons when all three types/subtypes co-circulated, i.e., 2014-2015, 2015-2016, and 2019-2020, individuals aged 5-14 years were the most affected by A/H3N2. On the other side, in all the analyzed seasons, the most affected age group by influenza B is 5-14 years with attack rates as high as 56.4% (95%CI: 47.1%-65.6%) during the 2012-2013 season. The elderly appears to be the least affected segment of the population across all types/subtypes and seasons, showing an attack rate consistently lower than 8.6%.

We estimate the effective reproduction number to be in the range 1.09-1.54 (see Tab. 1), in agreement with the literature on seasonal influenza [30]. We find that the reproduction number of A/H1N1pdm09 was highest in 2017-2018 season and reached the value of 1.4 (95%CI: 1.36-1.44), lower than values observed in Italy for the pandemic [5]. Only during two seasons, and for one influenza type/subtype per season only, the mean effective reproduction number exceeded 1.4: for A/H3N2 subtype in the 2011-2012 season (estimated mean 1.54, 95%CI: 1.33-1.85) and for B virus in the 2012-2013 season (estimated mean 1.48, 95%CI: 1.37-1.63).

**Table 1.**
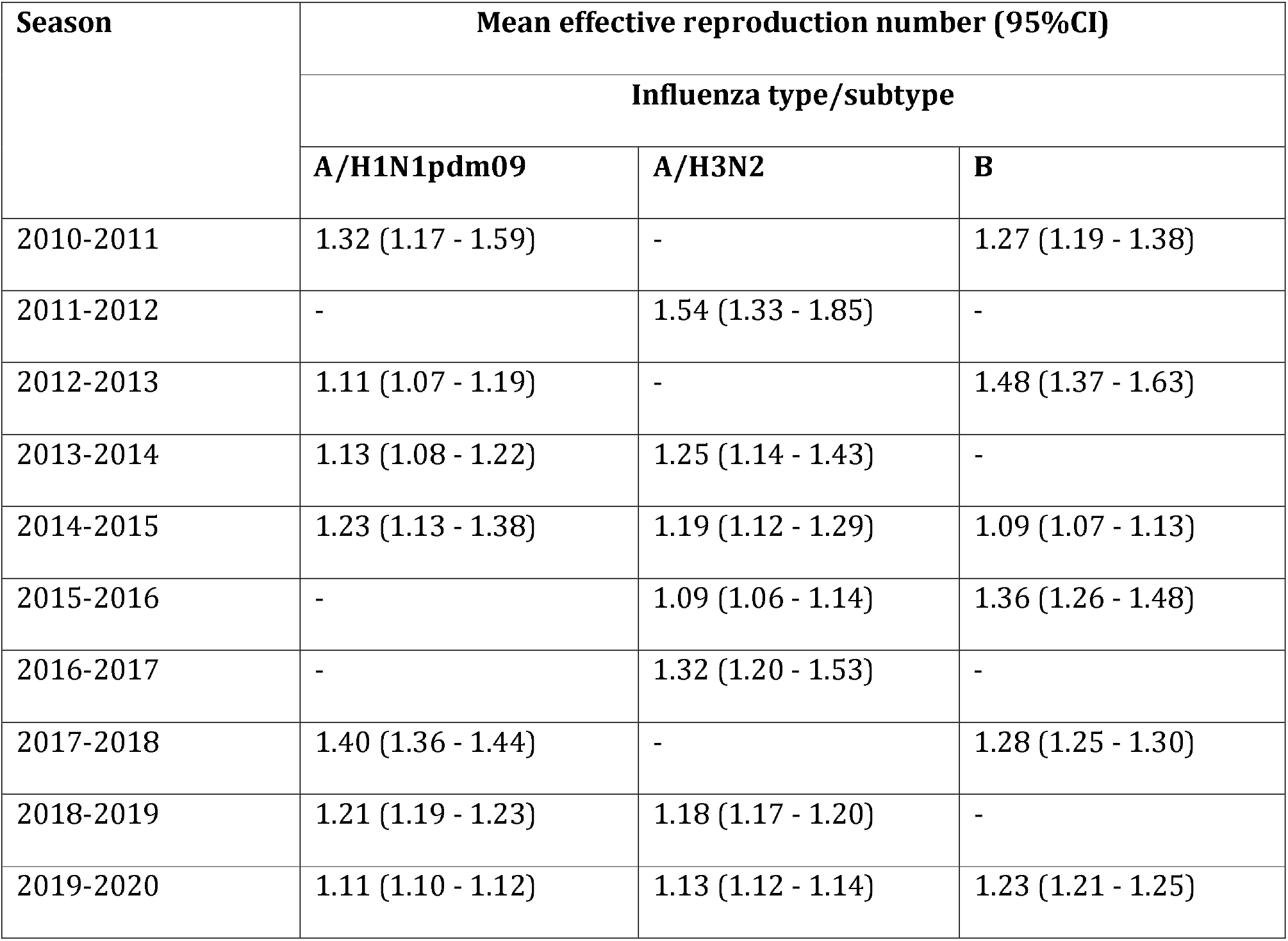
Estimated posterior distribution of the effective reproduction number (mean and 95%CI). Only types/subtypes that accounted for more than 15% of the seropositive samples are considered.

We estimate the susceptibility to infection to be rather constant across different seasons, but rather heterogeneous across different types/subtypes, suggesting that this epidemiological parameter is highly characteristic of each type/subtype (Fig. 3). In all seasons, we estimate the susceptibility to A/H1N1pdm09 infection to be in the range 0.19-0.38 for both 5-14 years and 15-64 years age groups, markedly different from what was estimated for the 2009 pandemic where school-age individuals were highly susceptible to infection [7,16,25,26,27,31].

**Figure 3.**
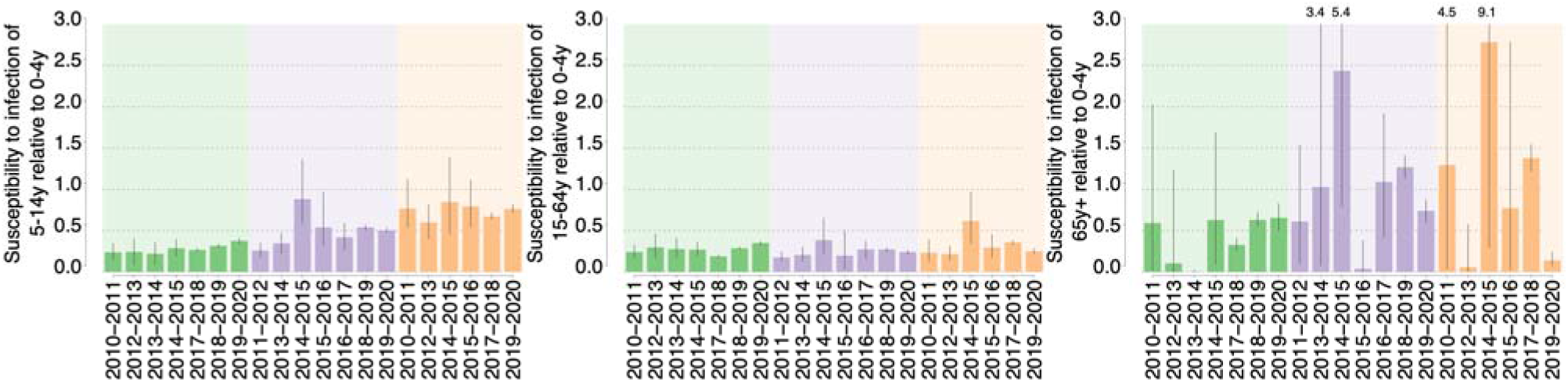
Estimated posterior distributions of the susceptibility to infection by age group (mean and 95%CI) relative to the 0-4 years age group (for which the susceptibility to infection is set to the reference value of 1). Only types/subtypes that accounted for more than 15% of the positive samples are considered. The values reported above the vertical lines in the right panels represent the 97.5% percentile of the distribution, when the value exceed the limit of the vertical axis.

A markedly larger susceptibility to infection was estimated for individuals aged 5-14 years for the other two types/subtypes, reaching during the 2014-2015 season values of 0.85 (95%CI: 0.46-1.39) for B influenza and 0.88 (95%CI: 0.58-1.37) for A/H3N2 influenza. For all types/subtypes, a highly variable susceptibility to infection is estimated for the elderly, with mean values ranging from nearly 0 up to 2.43. This is due to the low number of samples collected (and also testing positive) among ILI cases aged 65+ years, which results in highly variable Bayesian estimates. However, in the seasons where influenza circulation among the elderly is detected, the elderly tends to be more susceptible to A/H3N2 and B infection than to A/H1N1pdm09 infection.

## DISCUSSION

By analyzing epidemiological and surveillance data as well as seroepidemiological, socio-demographic, and contact data, we used mathematical modeling to characterize influenza transmission patterns over ten influenza seasons in Italy. We witnessed an alternation of a predominant type/subtype and the co-circulation of multiple types/subtypes. Nonetheless, we estimate the effective reproduction number to be mostly in the range 1.1-1.3 and rarely above 1.4, in general agreement with the literature [30]. Age-specific susceptibility to infection appears to be type/subtype-related, with a markedly larger susceptibility of 5-14 years old individuals to A/H3N2 and B influenza infection than to A/H1N1pdm09 influenza infection. Susceptibility to infection estimates for the elderly are highly variable both by type/subtype and by season. The estimated reporting rate to the Italian national surveillance system of about 1 case out of 3-5 is in line with the estimates found in previous studies [5,25,32].

The estimated total infection attack rate varied between about 18% to about 36% across seasons. When analyzing the infection attack rate by age, an even larger variability was estimated, mainly associated with the different infection patterns of the three types/subtypes. In fact, individuals aged 0-4 years are generally the most affected group by both influenza A subtypes while B influenza virus affects mainly individuals aged 5-14 years. However, A/H1N1pdm09 and B types/subtypes affect the elderly less than A/H3N2, probably due to the high degree of antigenic drift of the A/H3N2 virus [33], and the consequent lower vaccine effectiveness against A/H3N2 infection [20] and higher susceptibility to infection. Overall, individuals aged 14 years or less are the most affected segment of the population, coherently with seroepidemiological studies performed on the 2009 pandemic [16,34-36], probably due to both a larger susceptibility to infection, as measured in this study and in the literature [7,16,26,27], and to the larger number of social contacts of school-age individuals with respect to the rest of the population [23-25].

We estimate the elderly to be the least affected age group. However, this segment of the population is associated in the literature with larger hospitalization and mortality rates [37,38]. Therefore, also in the light of the rapid increase in the aging of populations throughout the world, this makes the study of immunosenescence in older individuals and the development of more immunogenic vaccines, two pipeline priorities in the control and prevention of influenza [39]. Moreover, although our estimates of the age-specific susceptibility to infection cannot be used to discern between biological, social, or hygienic determinants, they provide relevant indications for interventions targeting specific age segments of the population, school-age individuals above all.

These results support the importance of recommending the vaccine also to school-age healthy children, as already done by some EU countries and internationally [40], and call for further studies on school closure strategies as a possible non-pharmaceutical intervention to mitigate influenza spread in case of severe and/or pandemic seasons [25,41-46].

It is important to stress that our study suffer of some limitations. First, we do not consider the potential cross-protection among different types/subtypes and we do not model specifically co-circulation, as a more detailed serological data collection on levels of immunity for different influenza type/subtype of the Italian population in different age classes would be required. Therefore, instead of including several assumptions on the level of cross-protection induced by the infection with an influenza type/subtype to another type/subtype, we decided to consider the easiest assumption of no cross protection. Nonetheless, we would like to stress that this assumption does not affect our estimates of the total and age-specific infection attack rate. A second limitation is that we apply the estimated reporting rate to the surveillance system to all the analyzed seasons and types/subtypes. The calculation of type/subtype-and season-specific reporting rates was not possible due to the unavailability of the necessary data. However, our assumption is partially backed up by the findings of Carrat and colleagues [4] who estimated a similar share of individuals developing clinical symptoms after influenza infection for A/H1N1, A/H3N2, and B influenza types/subtypes.

## CONCLUSIONS

Our study provides indications on age- and type/subtype-specific incidence rates and susceptibility to infection as well as pathogen transmissibility, thus contributing to define a clear picture of the epidemiology of seasonal influenza in Italy. Our work provides relevant insights on age-specific targets for influenza immunization and non-pharmaceutical intervention plans, possibly also tailored on the circulating influenza type/subtype.

## Data Availability

The datasets analysed during the current study are available from the corresponding author on reasonable request.

## LIST OF ABBREVIATIONS

ILI: Influenza-like illness
GP: General practitioner
PD: Pediatrician
CRF: Case report form
NIC: National influenza center
AR: Attack rate
MCMC: Markov Chain Monte Carlo
NGM: Next Generation Matrix
Re: Effective reproduction number

## DECLARATIONS

### Ethics approval and consent to participate

Not appplicable

### Consent for publication

Not appplicable

### Availability of data and material

The datasets analysed during the current study are partly publicly available and partly available from the corresponding author on reasonable request.

### Author’s contribution

Study design: FT, SM, MA. Acquisition of data: EP, AB, GD, LC, CR. Data analysis: FT. Interpretation of results: FT, CR, SM, MA. Drafting of the manuscript: FT, MA. Critical revision of the manuscript: EP, AB, CR, SM. All authors have read and approved the final manuscript.

## Acknowledgments

Not applicable.

## Additional files

**Additional file 1: Supporting_information.pdf. Additional results**

