## Additional File 1 for "Characterizing the transmission patterns of seasonal influenza in Italy: lessons from the last decade"

### Appendix

#### Contents

1. Vaccination coverage
2. Virological data
3. Estimation of ILI reporting rate
4. Estimation of age-specific attack rates
5. Age-structured influenza transmission model
6. Bayesian analysis
7. References

#### 1. Vaccination coverage

Initial condition on the fraction of susceptible individuals at the beginning of each season are set by considering the observed age-specific vaccination coverage (see Tab. S1) and strain-specific vaccine effectiveness (VE) (see Tab S2).

**Tab S1.** Seasonal influenza vaccine coverage by age group (m=month, y=year) for influenza vaccine in the period 2010-2020, as reported to the Italian National Institute of Health (1).

| SEASON | 6-23m | 2-4y | 5-8y | 9-14y | 15-17y | 18-44y | 45-64y | ≥65y | Total |
|---|---|---|---|---|---|---|---|---|---|
| 2010-2011 | 2.9% | 4.4% | 4.2% | 3.7% | 3.5% | 3.5% | 11.5% | 60.2% | 17.4% |
| 2011-2012 | 2.2% | 4.2% | 4.5% | 3.4% | 3.6% | 3.4% | 12.1% | 62.7% | 18.2% |
| 2012-2013 | 1.5% | 2.6% | 2.6% | 2.0% | 2.1% | 2.1% | 9.0% | 54.2% | 15.0% |
| 2013-2014 | 1.3% | 2.5% | 2.6% | 2.1% | 2.3% | 2.5% | 9.5% | 55.5% | 15.8% |
| 2014-2015 | 1.1% | 1.8% | 1.9% | 1.5% | 1.5% | 1.9% | 7.5% | 48.7% | 13.6% |
| 2015-2016 | 1.1% | 1.8% | 1.8% | 1.4% | 1.6% | 1.8% | 7.7% | 49.9% | 14.1% |
| 2016-2017 | 1.5% | 2.6% | 2.4% | 1.8% | 1.9% | 2.2% | 8.5% | 52.0% | 15.1% |
| 2017-2018 | 1.4% | 2.4% | 2.2% | 1.8% | 2.5% | 2.2% | 8.7% | 52.7% | 15.4% |
| 2018-2019 | 1.7% | 3.1% | 2.5% | 1.8% | 2.2% | 2.6% | 8.9% | 53.1% | 15.9% |
| 2019-2020 | 1.7% | 3.1% | 2.5% | 1.8% | 2.2% | 2.6% | 8.9% | 53.1% | 16.2% |

**Tab S2.** Pooled VE estimates by strain, accounting for reported antigenic similarity of A/H3N2 viruses to the vaccine strain as found in (2).

| Strain | A/H1N1pdm09 | B | A/H3N2 similar* | A/H3N2 variant* |
|---|---|---|---|---|
| Vaccine effectiveness | 61.3% | 52.3% | 33.0% | 22.0% |

*Similar and variant A/H3N2 refer to antigenically matched or mismatched A/H3N2 viruses, respectively. For the 2014/15 and 2016/17 we used "variant" VE, as there is evidence of mismatch between the vaccine and the circulating strain (3), while "similar" VE is used for the other seasons.

#### 2. Virological data

The share of samples collected among ILI cases testing positive to each strain and total number of analyzed samples are obtained from data collected in Lombardy region (4) – data reported in Tab S3.

**Tab S3.** Circulating strains and relative share of samples testing positive to each strain (4) over the period 2010-2020. We highlighted in blue the strains that above the 15% threshold that we used to conduct the Bayesian analysis.

| Season | A/H1N1pmd09 | A/H3N2 | B |
|---|---|---|---|
| 2010-2011 | **60.6%** | 4.1% | **35.3%** |
| 2011-2012 | 0.4% | **87.0%** | 12.6% |
| 2012-2013 | **18.6%** | 3.8% | **77.6%** |
| 2013-2014 | **30.7%** | **69.3%** | 0.0% |
| 2014-2015 | **43.0%** | **35.0%** | **22.0%** |
| 2015-2016 | 12.2% | **16.9%** | **70.9%** |

| | | | |
|---|---|---|---|
| 2016-2017 | 0.4% | **94.4%** | 5.2% |
| 2017-2018 | **37.2%** | 2.3% | **60.5%** |
| 2018-2019 | **50.0%** | **50.0%** | 0.1% |
| 2019-2020 | **26.8%** | **40.2%** | **33.0%** |

In this study, we consider the circulation of a strain to be negligible during a season if the share of samples testing positive to that strain below 15%. All Influenza strains in all seasons are included in the analysis of the infection attack rate, while only influenza strains that are above a 15% threshold are considered in the Bayesian analysis.

The shares of *ILI* samples by age group $a$ testing positive for strain $s$ in season $y$, $f_a(y,s)$, are also obtained from the virological surveillance database of Lombardy region (4) – data reported in Tab S4.

**Tab S4.** Shares of ILI samples testing positive for the three strains on the overall number of tested samples in each age class (0-4y, 5-14y, 15-64y and 65+y). Data obtained from virological surveillance conducted in Lombardy region (4) until 2016-2017 and from the national virological surveillance in the last three seasons.

| SEASON | A/H1N1pdm09 | A/H3N2 | B |
|---|---|---|---|
| **2010-2011** | 0.28; 0.12; 0.3; 0.15 | | 0.13; 0.33; 0.11; 0.13 |
| **2011-2012** | | 0.44; 0.41; 0.38; 0.42 | |
| **2012-2013** | 0.08; 0.04; 0.10; 0.04 | | 0.31; 0.55; 0.27; 0.07 |
| **2013-2014** | 0.13; 0.07; 0.13; 0.04 | 0.29; 0.3; 0.21; 0.26 | |
| **2014-2015** | 0.26; 0.16; 0.25; 0.17 | 0.11; 0.32; 0.14; 0.27 | 0.04; 0.13; 0.13; 0.12 |
| **2015-2016** | | 0.07; 0.11; 0.05; 0.11 | 0.21; 0.55; 0.26; 0.16 |
| **2016-2017** | | 0.37; 0.57; 0.47; 0.5 | |
| **2017-2018** | 0,22; 0,18; 0,15; 0,05 | – | 0,14; 0,38; 0,22; 0,20 |
| **2018-2019** | 0,18; 0,17; 0,21; 0,11 | 0,13; 0,29; 0,13; 0,19 | – |
| **2019-2020** | 0,07; 0,06; 0,08; 0,04 | 0,11; 0,19; 0,08; 0,07 | 0,09; 0,31; 0,07; 0,01 |

As validation of the virological surveillance data reported to Lombardy region, we compare the data for all age groups combined together as reported to Lombardy virological surveillance to those reported to the Italian national virological surveillance for all the seasons from 2009-2010 to 2016-2017 (see Fig S1). We find a significant correlation between the two dataset both overall and for each strain: all strains, Pearson correlation: 0.97, p-value <0.0001; A/H1N1pdm09, Pearson correlation: 0.98, p-value <0.0001; A/H3N2, Pearson correlation: 0.99, p-value <0.0001; B, Pearson correlation: 0.99, p-value <0.0001.

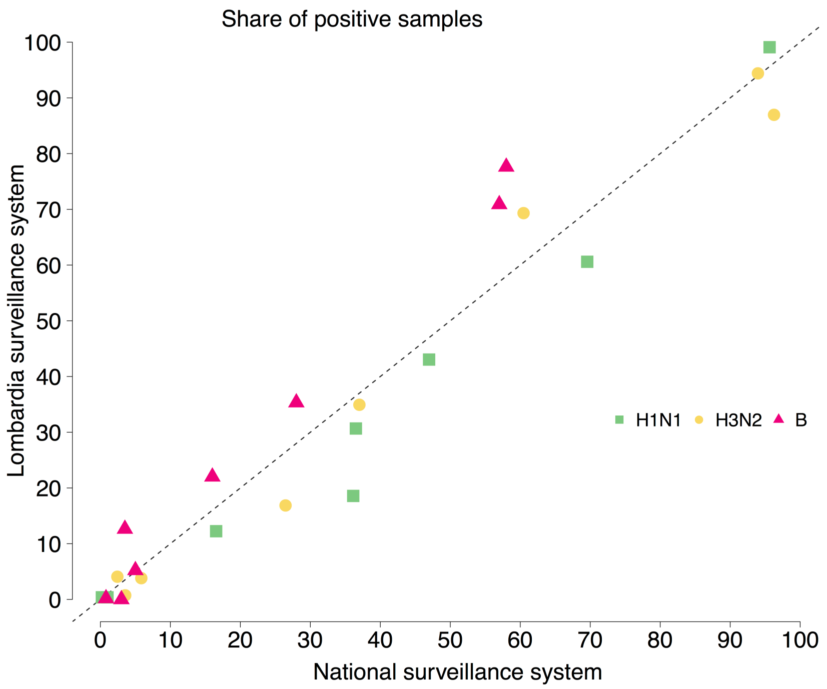


**Fig. S1.** Scatterplot of the share of samples testing positive for each strain in the seasons from 2009-2010 to 2016-2017, as reported to the Italian national virological surveillance system (here denoted as National surveillance system) (3) and to Lombardy region virological surveillance systems (here denoted as Lombardy surveillance system) (4).

#### 3. Estimation of ILI reporting rate

We consider, as a starting point, levels of immunity of the Italian population in different age classes before and after the A/H1N1 influenza pandemic in 2009-2010 (5,6). The two sero-epidemiological studies are based on 1,152 and 1,436 leftover sera collected among individuals of the general population before and right after the end of the 2009 influenza pandemic, respectively.

Using these two datasets we estimate the infection attack rates of the 2009 influenza pandemic for two age groups: 0-14y and 15y+ (denoted as $AR_{0-14}$ and $AR_{15+}$, respectively), as follows:

$$AR_a = (POS_a^{post} - POS_a^{pre})$$

where:

- $a$ denotes the age class;
- $\mathrm{POS}_a^{post}$ and $\mathrm{POS}_a^{pre}$ are the shares of samples testing positive to A/H1N1 A/California/07/2009 (A/H1N1pdm09) in age group $a$ before and after the 2009-10 pandemic, respectively.

The attack rate detected by the Italian surveillance system in the same age groups (denoted as $AR_{0-14}^{ILI}$ and $AR_{15+}^{ILI}$ for the 0-14y and 15+y age groups, respectively), can be computed as follows:

$$AR_a^{ILI} = f_{H1N1}^{+} * \sum_{w=1}^{32} ILI_a(w)$$

where:

- $f_{\mathrm{H1N1}}^{+}$ is the the share of samples collected among ILI cases testing positive to A/H1N1pdm09;
- $a$ denotes the age class;
- $w$ denotes the surveillance week;

- $ILI_a(w)$ is the incidence of ILI cases reported to the Italian epidemiological influenza surveillance system (7) during the pandemic for age group $a$ on week $w$.

Note that during the 2009-2010 influenza season, 96.4% of samples collected among ILI cases testing positive for influenza infection, tested positive to A/H1N1pdm09 infection (3).

The reporting rates of the surveillance system for the age group 0-14 years and 15+ years, (denoted as $r_{0-14}$ and $r_{15+}$, respectively) can be estimated by the following equation:

$$r_a = \frac{AR_a^{ILI}}{AR_a}$$

Essentially, $r_{0-14}$ and $r_{15+}$ represent the share of influenza infections that are detected by the Italian influenza surveillance system. In the rest of the analysis, we assume these reporting rates are the same for other types and subtypes of influenza virus and for each season. Although this is a strong assumption, we find similar estimates of the reporting rate for other surveillance systems estimated for different influenza seasons after the 2009 influenza pandemic (8,9).

#### 4. Estimation of age-specific infection attack rates

For each analyzed influenza season (from 2010-11 to 2019-20), the age-specific infection attack rates are calculated by combining the reporting rates mentioned above with the observed ILI incidence in the different age classes (0-4, 5-14, 15-64, and 65+ years) and virological data by age. In particular, we use the following equation:

$$AR_s^a(y) = \frac{\sum_{w=1}^{28} ILI_a(w;y) * f_s^a(y)}{r_a}$$

where:

- $a$ is the age class;
- $s$ is the strain;
- w is the surveillance week;
- $y$ is the season;
- $ILI_a(w;y)$ is the incidence of $ILI$ cases reported to the Italian surveillance system (7) for age group $a$ on week $w$ of season $y$;
- $f_s^a$(y) is the share of ILI samples for age group $a$ testing positive for strain $s$ in season $y$;
- $r_a$ is the reporting rate for age group $_a$ (see previous section).

Note that, as for the reporting rate we have only two age groups (0-14 and 15+ years) while for the ILI cases we have four age groups (0-4, 5-14, 15-64, and 65+ years), for the age groups 0-4 and 5-14 we apply $r_{0-14}$ and for age groups 15-64 and 65+ we apply $r_{15+}$.

#### 5. Age-structured influenza transmission model

Influenza transmission for the three considered strains is simulated through a deterministic non-stationary age-structured SIER model (6) stratified in 85-years age classes and based on the assumption of heterogeneous mixing by age. The epidemiological transitions for each individual's age are described by the following system of ordinary differential equations:

$$\begin{cases} \frac{dS(a,t)}{dt} = -\beta\rho_a S(a,t) \sum_{a'=0}^{85} \frac{C_{a;a'} I(a',t)}{N(a',t)} \\ \frac{dE(a,t)}{dt} = \beta\rho_a S(a,t) \sum_{a'=0}^{85} \frac{C_{a;a'} I(a',t)}{N(a',t)} - \delta E(a,t) \\ \frac{dI(a,t)}{dt} = \delta E(a,t) - \gamma I(a,t) \end{cases}$$

where:

- $t$ represents the time;
- *S(a,t)*, *E(a,t)*, *I(a,t)* and *R(a,t)* represent the number of susceptible, latent, infectious and removed individual of age $a$ at time *t*, respectively;
- *N(a,t)=S(a,t)+E(a,t)+I(a,t)+R(a,t)* represents the total population of age $a$ at time *t;*
- $\beta$ is the transmission rate;
- $\rho_a$ denotes the susceptibility to infection of individuals of age $a$ relative to the susceptibility to infection of individuals aged 0-4 years that is set to 1;
- $C$ is contact matrix by age, where each element $C_{a;a'}$ represents the average number of contact between individuals of age $a$ and individuals of age $a'$;
- $1/\delta$ is the average duration of the latent period;
- $1/\gamma$ is the average duration of the infectious period.

We initialized the model with 10 infectious individuals (and zero latent individual) at the beginning of each season, distributed randomly across different ages. The initial numbers of susceptible and removed individuals account for the observed age-specific vaccination coverage and strain-specific vaccine effectiveness and are computed as follows:

$$S(0) = (N(0) - \epsilon(s)) \sum_{a'=0}^{85} \delta(a,a')c(a',0)N(a',0) - 10$$

$$R(0) = \epsilon(s) \sum_{a'=0}^{85} \delta(a,a')c(a',0)N(a',0) + 10$$

where:

- $S(0)$ and $R(0)$ are respectively the initial numbers of susceptible and removed individuals in the population (irrespectively of the age) at the beginning of each season;
- $N(0)$ is the total population at the beginning of each season;
- $c(a,0)$ is the coverage for individuals of age $a$ at the beginning of each season (see Sec. S1);
- $\epsilon(s)$ represents the strain-specific vaccine effectiveness (see Sec. S1);
- $\delta(a,a')$ is the Dirac delta function, equal to 1 if $a = a'$ and 0 otherwise.

The contact matrix for the Italian population is taken from (10). The average duration of the latent period ($1/\delta$) is set to 1.5 days (11) and the infectious period ($1/\gamma$) is set to 1.2 days in such a way that the resulting generation (12) is 2.7 days, in agreement with influenza literature (13).

#### 6. Bayesian analysis

We calibrate the influenza transmission model (see Sec. 5) separately for each season and circulating strain. For each season and strain, we have four free parameters (the transmission rate, $\beta$, and the susceptibilities to infection $\rho_{1,\ldots 3}$ of individuals in age classes 5-14, 15-64 and >64 years, respectively) whose posterior distributions are estimated through a Bayesian approach. In particular, model calibration is carried out by means of a differential evolution Markov chain Monte Carlo sampling (14) applied to the binomial likelihood of the season- and strain- specific influenza infection attack rates $AR_s^a(y)$. In particular, for each season $y$, the likelihood of the transmission rate and susceptibility to infection by age group given the infection attack rates by age is defined as:

$$\mathcal{L}\left(\beta, \rho_{1,\dots 3} \middle| n, AR_s^a(y)\right) =$$

$$= \prod_{a=1}^{4} \binom{n(a)}{n(a)AR_s^a(y))} p(a;\beta,\rho_a)^{n(a)AR_s^a(y)} \left(1 - p(a;\beta,\rho_a)\right)^{n(a)(1-AR_s^a(y))}$$

where:

- $a$ runs over the four age classes for which the infection attack rates is known (i.e., 0-4, 5-14, 15-64, and 65+ years);
- n(a) is the number of individuals in the age class $a$ tested for a specific strain during season-specific virological studies;
- $AR_s^a(y)$ is the age-specific infection attack rates for strain $s$ during season $y$ (see Sec. S4);
- $p(a;\beta,\rho_a)$ is the estimated infection attack rate in age class $a$ as obtained by model simulation with a specific transmission rate β and susceptibility to infection $\rho_{1,\dots 3}$.

For each influenza season and strain, we run five chains of 50,000 iterations using different starting points for β and $\rho_{1,\dots 3}$. For β and $\rho_{1,\dots 3}$ we use the same prior distribution: a flat distribution between 0 and 1,000. MCMC convergence and the length of the burn-in period are assessed by checking via visual inspection of the trace plots associated with the different chains, i.e. the sequence of accepted parameter values are approximately characterized by a constant width and average, therefore proving good mixing of the parameters.

As an example, we show the trace plots associated with one chain for the 2010-11 season and A/H1N1pdm09 strain is shown in Fig. S2 (panels A-D), along with the estimated infection attack rate by age (panel E) and posterior distributions of $R_{eff}$ and susceptibility to infection by age group (panels F and G).

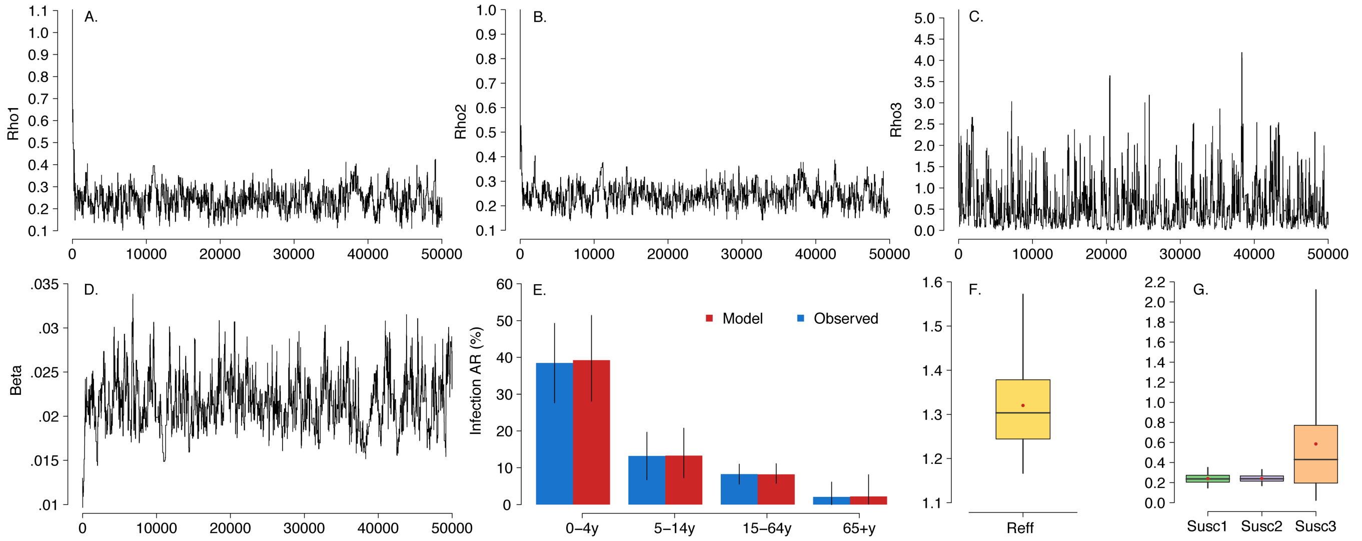


**Fig. S2.** Model calibration for A/H1N1pdm09 during the 2010-2011 influenza season. **A** Trace plot of the susceptibility to infection of the 5-14y age group relative to the 0-4y age group. **B** As A, but for the 15-64y age group. **C** As A, but for the 65+y age group. **D** As A, but for the transmission rate. **E** Observed mean age-specific infection attack rates (see Sec. 4) and estimated by the calibrated influenza transmission model. Vertical lines represent 95% credible intervals. **F** Posterior distribution (quantiles: 0.025, 0.25, 0.5, 0.75, and 0.975) of the effective reproduction number. **G** Posterior distribution (quantiles: 0.025, 0.25, 0.5, 0.75, and 0.975) of the susceptibilities to infection for the 5-14y (Susc1), 15-64y (Susc2), and 65+y (Susc3) age groups relative to the 0-4y age group.
